# Waning protection of long-acting RSV monoclonal antibodies in infants: a Bayesian analysis of clesrovimab and nirsevimab trial data

**DOI:** 10.64898/2026.06.15.26355703

**Authors:** Dexin Gong, Stefan Flasche, David Hodgson

**Author notes:** Corresponding author: Dexin Gong.

## Abstract

Clesrovimab and nirsevimab are long-acting monoclonal antibodies used to prevent respiratory syncytial virus (RSV) disease in infants, but waning protection in the first year of life is incompletely characterised. We applied a published Bayesian inference framework to clesrovimab and pooled nirsevimab trial data to estimate time-varying efficacy against medically attended RSV lower respiratory tract infection (LRTI) and RSV-associated hospitalisation, accounting for differences in placebo-arm event timing between trials. Estimated clesrovimab efficacy declined from 60.7% (95% CrI: 46.3–72.6) shortly after dosing to 38.3% (8.6–52.9) at six months against medically attended RSV LRTI, and from 87.1% (71.2–96.2) to 49.6% (10.4–70.7) against RSV-associated hospitalisation. For nirsevimab, corresponding estimates declined from 86.9% (75.4–95.0) to 53.8% (27.4–69.7) against LRTI, and from 77.5% (52.6–91.8) to 49.7% (15.7–68.3) against hospitalisation. After accounting for differences in RSV exposure timing and LRTI endpoint definitions between trials, we found no evidence of a difference in efficacy or waning between clesrovimab and nirsevimab.

## 1. Introduction

Respiratory syncytial virus (RSV) is a leading cause of acute lower respiratory tract infection (LRTI) in infants, responsible for an estimated 33.0 million episodes and 3.6 million hospitalisations per year in children under five worldwide [1]. Two long-acting monoclonal antibodies (mAbs) targeting the RSV fusion protein, nirsevimab and clesrovimab, are now licensed for infant protection. Trials established efficacy for both products with observation periods spanning 150 days post-dose [2,3], albeit with exposure concentrated early in that period.

For nirsevimab, post-licensure effectiveness studies and pragmatic trial data have begun to characterise protection beyond the 150-day trial window [4–6]. Yet, equivalent evidence for clesrovimab beyond the 150-day trial window and the waning within the first 150 days is not yet available. Waning assumptions therefore remain a sensitive input for programme design, particularly in settings with prolonged or year-round RSV circulation [7].

No head-to-head trial has compared clesrovimab and nirsevimab, and published cumulative efficacy estimates are difficult to compare directly because trial conditions, including RSV exposure timing with respect to immunisation, differed across programmes. We used a previously developed Bayesian inference framework [8] to publicly available aggregate data from the CLEVER (clesrovimab) and pooled phase 2b/MELODY (nirsevimab) trials [2,3] to derive comparable waning estimates.

## 2. Methods

We analysed publicly available aggregate data from the CLEVER phase 2b–3 trial of clesrovimab [3] and the pooled phase 2b/MELODY analysis of nirsevimab [2], giving four product–endpoint combinations: medically attended RSV LRTI and RSV-associated hospitalisation for each product. Published Kaplan– Meier curves were digitised together with arm-specific event counts and person-time. Observed event timing was summarised in 30-day intervals after dosing and stratified by trial arm. Trial-reported 0–150-day cumulative efficacy estimates were retained for comparison with model-derived estimates. Estimates beyond 150 days were not summarised as cumulative outcomes given the absence of observed post-trial RSV exposure data.

We used a previously described Bayesian waning framework [8]. Within each trial, placebo and intervention arms were assumed to share a common time-varying force of RSV exposure, modelled with a Gaussian process prior informed by placebo-arm event data. Protection in the intervention arm was modelled as a time-varying multiplicative reduction in the force of infection. Waning was parameterised using three candidate specifications: exponential, Erlang-2, and Erlang-3. Candidate specifications were compared using leave-one-out information criterion (LOOIC) [9]. The primary analysis used the Erlang-3 specification across all four product–endpoint combinations, with exponential and Erlang-2 as sensitivity analyses. Scale-parameter upper bounds of 365, 180, and 120 days for the exponential, Erlang-2, and Erlang-3 specifications, respectively, bounded the implied mean waning duration above by approximately one year (since the Erlang-k mean equals k times the scale parameter), avoiding weakly identified long-duration waning from a 0–150-day trial window.

Posterior inference was performed using Hamiltonian Monte Carlo, with four chains and 1,000 warm-up and 1,000 sampling iterations each. Convergence was assessed by R-hat diagnostics and visual inspection of trace plots. Efficacy was reported at specific post-dose time points as posterior medians with 95% credible intervals. The six-month estimate refers to the time-specific model estimate at day 180 and is not a cumulative efficacy. For each posterior draw, model-derived 0–150-day cumulative estimates were calculated as the placebo-risk-weighted average of the time-specific efficacy curve, with weights given by the draw-specific expected number of placebo events in each interval. These estimates were compared with trial-reported 0–150-day cumulative efficacy as internal consistency checks. Reconstructed placebo-incidence curves were compared with observed interval-specific rates as an additional model check. As an exploratory sensitivity analysis, the clesrovimab medically attended RSV LRTI model was additionally refitted to digitised Kaplan–Meier data for the CLEVER post hoc endpoint more closely aligned with the nirsevimab case definition.

This analysis used only publicly available aggregate data and did not require additional ethical approval. Analyses were conducted in R [10] using CmdStanR [11]. The analysis code is available at https://github.com/dexinGONG/rsv-mab-waning.

## 3. Results

All primary models converged satisfactorily, with R-hat below 1.01 for all parameters (Supplementary Figures S3 and S4). LOOIC differences across the three waning specifications were small (Supplementary Table S1). We therefore retained Erlang-3 as the primary specification across all four product–endpoint combinations for comparability, with exponential and Erlang-2 reported as sensitivity analyses. Reconstructed placebo-group incidence broadly captured the observed pattern over the 0–150-day trial window, with some divergence at late time points for the clesrovimab fits (Supplementary Figure S1). Under the primary Erlang-3 specification, model-estimated clesrovimab efficacy declined from 60.7% (95% CrI: 46.3–72.6) shortly after administration to 38.3% (8.6–52.9) at six months against medically attended RSV LRTI under the CLEVER primary endpoint definition, and from 87.1% (71.2–96.2) to 49.6% (10.4–70.7) against RSV-associated hospitalisation. For nirsevimab, corresponding estimates declined from 86.9% (75.4–95.0) to 53.8% (27.4–69.7) against LRTI, and from 77.5% (52.6–91.8) to 49.7% (15.7–68.3) against hospitalisation (Figure 1A–B; Table 1). Modelled early efficacy (before day 90) against medically attended RSV LRTI appeared lower for clesrovimab than for nirsevimab (Figure 1A), whereas six-month estimates had overlapping uncertainty intervals.

**Table 1.** Model-estimated time-specific efficacy and 0–150-day cumulative efficacy.

| Endpoint and Product | Modelled initial efficacy* | Modelled efficacy at 6 months (day 180) | Modelled 0–150-day cumulative efficacy | Trial 0–150-day cumulative efficacy |
| --- | --- | --- | --- | --- |
| ***Medically attended RSV LRTI*** |  |  |  |  |
| Clesrovimab, primary endpoint | 60.7 (46.3–72.6) | 38.3 (8.6–52.9) | 57.0 (43.6–68.4) | 60.4 (44.1–71.9) |
| Nirsevimab | 86.9 (75.4–95.0) | 53.8 (27.4–69.7) | 80.0 (69.7–88.1) | 79.5 (65.9–87.7) |
| Clesrovimab, post hoc aligned endpoint† | 88.5 (78.1–95.3) | 60.7 (26.5–73.1) | 84.8 (74.9–92.2) | 88.0 (76.1–94.0) |
| ***RSV-associated hospitalisation*** |  |  |  |  |
| Clesrovimab | 87.1 (71.2–96.2) | 49.6 (10.4–70.7) | 80.5 (65.8–90.6) | 84.2 (66.6–92.6) |
| Nirsevimab | 77.5 (52.6–91.8) | 49.7 (15.7–68.3) | 71.2 (48.6–86.9) | 77.3 (50.3–89.7) |

\*Initial and six-month efficacy are posterior medians at day 0 and day 180 post-dose. Modelled 0–150-day cumulative estimates are placebo-risk-weighted posterior means computed using the model-estimated placebo event distribution. Trial estimates are reported 0–150-day cumulative efficacy. CrI, credible interval; CI, confidence interval. † Exploratory post hoc CLEVER endpoint aligned with the nirsevimab case definition (10 active vs. 42 placebo events through day 150); see Supplementary Figure S5.

**Figure 1.**
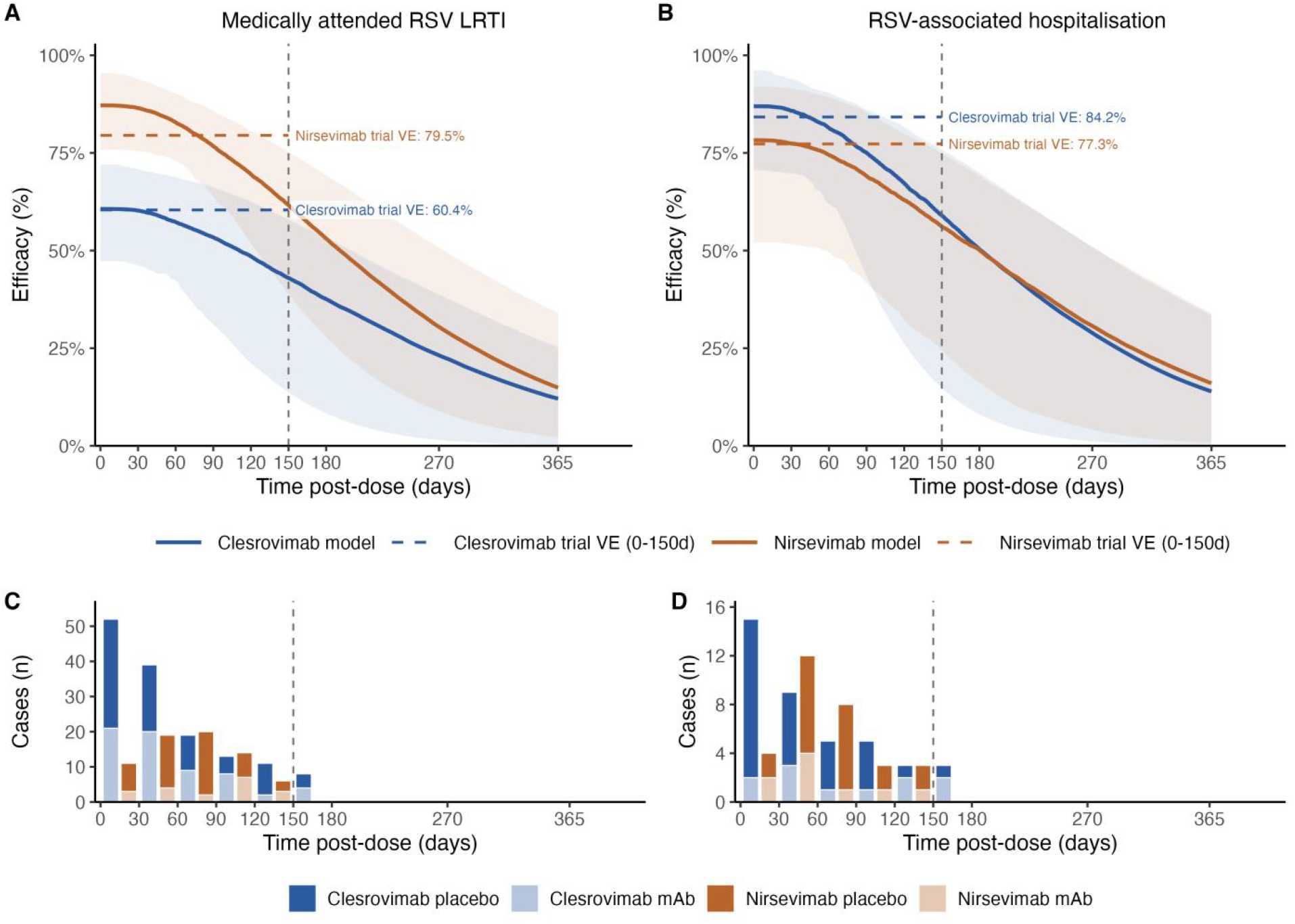
Model-estimated time-varying efficacy (panels A and B) and observed event timing (panels C and D) for clesrovimab and nirsevimab. Panels A and B show the estimated time-varying efficacy against medically attended RSV LRTI and RSV-associated hospitalisation. Solid lines show posterior median efficacy estimates and shaded areas show 95% credible intervals. Horizontal dashed lines show trial-reported 0–150-day cumulative efficacy as a reference. Panels C and D show reconstructed event counts by 30-day interval after dosing, stratified by trial arm for RSV LRTI and RSV-associated hospitalisation, respectively; the vertical dashed line marks the 150-day efficacy window.

Observed exposure differed between trials (Figure 1C–D). In CLEVER, placebo-arm events were concentrated soon after dosing, with 68% of medically attended RSV LRTI events and 68% of RSV-associated hospitalisation events occurring within 60 days. In the pooled phase 2b/MELODY analysis, events were shifted later, with 65% and 71% of corresponding events occurring between days 30 and 90, i.e. when some of the protective effect may have waned.

The model results remained qualitatively similar in all sensitivity analyses. Differences across alternative waning specifications were small (Supplementary Table S1; Supplementary Figure S2). Relaxing the Erlang-3 scale parameter upper bound had little effect on 0–150-day cumulative efficacy (Supplementary Table S2). The exploratory CLEVER post hoc endpoint-alignment analysis attenuated the apparent LRTI difference seen under the CLEVER primary endpoint (Table 1; Supplementary Figure S5).

## 4. Discussion

Applying a Bayesian efficacy-waning framework to reported illness-event timing from licensing trials, we estimated time-varying protection for clesrovimab and nirsevimab while accounting for differences in event timing between trials. Six-month time-specific efficacy estimates had overlapping uncertainty intervals across products for both endpoints, although uncertainty was wider for hospitalisation. In HARMONIE, nirsevimab efficacy against RSV-associated LRTI hospitalisation through 180 days was 82.7% (95% CI: 67.8–91.5) [5]. Cohort and case–control studies in the US have also reported high effectiveness against RSV-associated hospitalisation and severe outcomes during the first season [4,6]. These cumulative or season-long estimates are not directly comparable with the time-specific values reported here but are consistent with sustained early-season protection. Pharmacokinetic data support the biological plausibility of waning passive antibody-mediated protection. Nirsevimab and clesrovimab have antibody half-lives of approximately 69 days and approximately 44–45 days [12,13], although serum concentration may not translate linearly into clinical efficacy.

Endpoint definitions and ascertainment differed between the two trials. A post hoc CLEVER analysis using a case definition more closely aligned with the MELODY nirsevimab criteria yielded 88.0% efficacy (95% CI: 76.1–94.0) against medically attended RSV LRTI, compared with 60.4% under the CLEVER primary endpoint and 79.5% (65.9–87.7) in the pooled phase 2b/MELODY analysis [2,3]. Refitting our model to digitised data from this post hoc endpoint yielded clesrovimab estimates similar to those for nirsevimab (Table 1; Supplementary Figure S5), though based on fewer events. The lower apparent clesrovimab estimate against medically attended RSV LRTI (Figure 1A) therefore appears to be driven substantially by case-definition stringency rather than product-level efficacy alone. The direction of the apparent early difference was not consistent across endpoints, and hospitalisation estimates were imprecise, further cautioning against a product-level interpretation. For hospitalisation, residual differences in admission practices and ascertainment may add heterogeneity that cannot be quantified from aggregate data.

The analysis needs to be considered in light of some key limitations. Aggregate rather than individual-participant data precluded adjustment for covariates such as weight or RSV subtype, though the pooled estimates we replicated were broadly consistent with published subgroup-adjusted analyses. Endpoints were analysed independently and within-individual correlation was not modelled, which affects joint uncertainty but not the marginal time-varying efficacy estimates. Residual programme-level differences in populations, ascertainment, admission practices, and seasonality could not be fully addressed. The framework also requires placebo-arm time-to-event data and cannot be applied to post-licensure studies; however, our estimates for the nirsevimab trial are generally in line with post-marketing estimates.

## 5. Conclusion

Overall, we found no evidence that the efficacy or the rate of waning differed between nirsevimab and clesrovimab.

## Data Availability

All data used in this study were derived from publicly available aggregate clinical trial data. The extracted data, analysis code, and materials required to reproduce the analyses are available in the GitHub repository listed below. No individual-level participant data or identifiable information were used.

https://github.com/dexinGONG/rsv-mab-waning

## Declaration of generative AI and AI-assisted technologies in the writing process

During the preparation of this work, Dexin Gong used ChatGPT and Claude to support language editing and manuscript formatting. After using these tools, the authors reviewed and edited the content as needed and take full responsibility for the content of the publication.

## CRediT authorship contribution statement

Dexin Gong: Data curation, Formal analysis, Visualization, Writing – original draft, Writing – review and editing.

Stefan Flasche: Conceptualization, Methodology, Supervision, Writing – review and editing.

David Hodgson: Conceptualization, Methodology, Supervision, Writing – review and editing.

## Acknowledgements

The authors thank Masinde Augustine and Ayaka Monoi for helpful discussions and for sharing relevant background materials that informed the interpretation of this analysis. The authors also thank the investigators of the CLEVER, phase 2b, and MELODY trial programmes for making their results publicly available, which enabled this secondary modelling analysis.

## Data availability

The analysis used publicly available data from the referenced clinical trial reports. Digitised data and analysis code are publicly accessible at https://github.com/dexinGONG/rsv-mab-waning.

*All authors attest they meet the ICMJE criteria for authorship*.

