## Supplementary Information for "Waning protection of long-acting RSV monoclonal antibodies in infants: a Bayesian analysis of clesrovimab and nirsevimab trial data"

***Supplementary Figure S1***

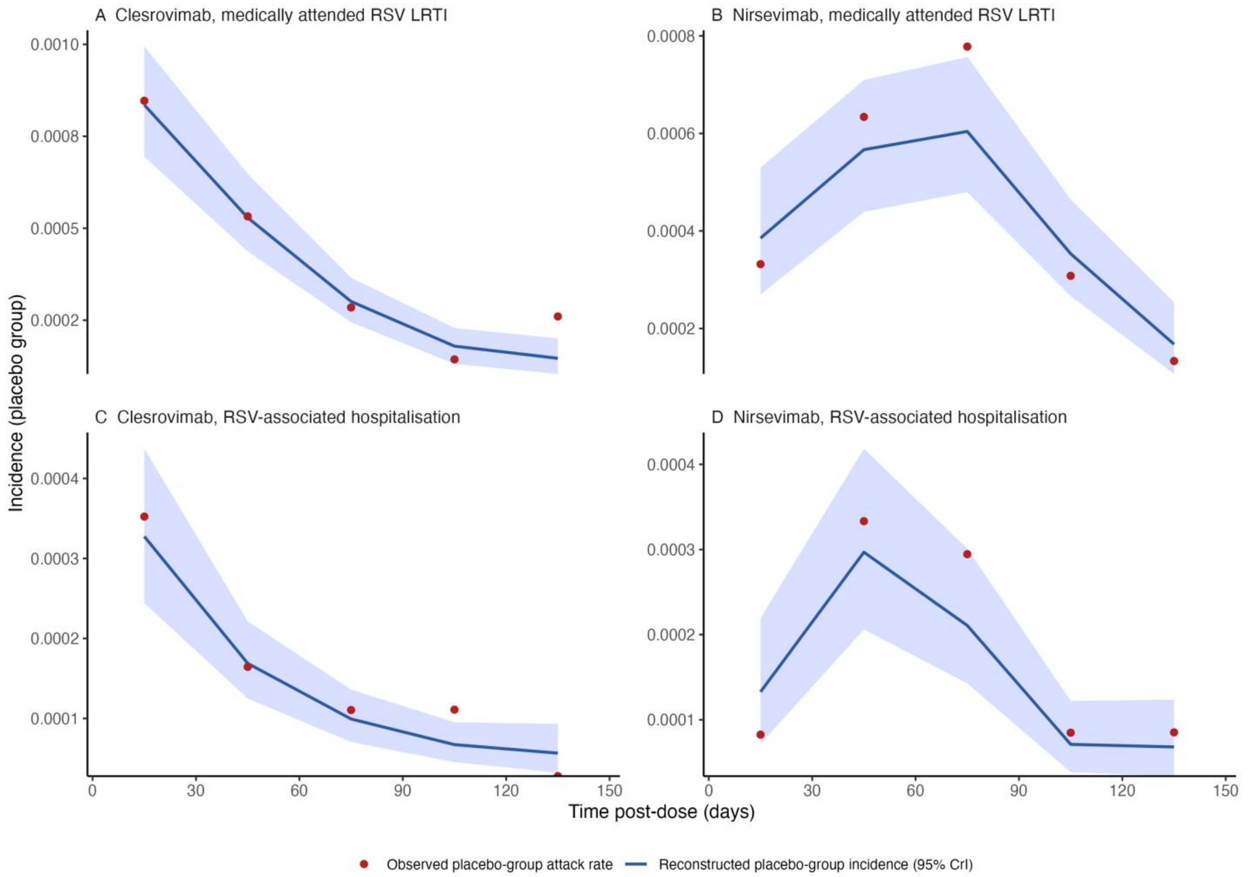


**Supplementary Figure S1.** Reconstruction of placebo-group incidence from digitised Kaplan–Meier data. Panels show placebo-group incidence for (A) clesrovimab against medically attended RSV lower respiratory tract infection (LRTI), (B) nirsevimab against medically attended RSV LRTI, (C) clesrovimab against RSV-associated hospitalisation, and (D) nirsevimab against RSV-associated hospitalisation. Solid lines represent posterior median reconstructed incidence, shaded areas represent time-specific 95% credible intervals, and red points represent observed interval-specific incidence rates derived from reconstructed event counts and person-time.

### ***Supplementary Figure S2***

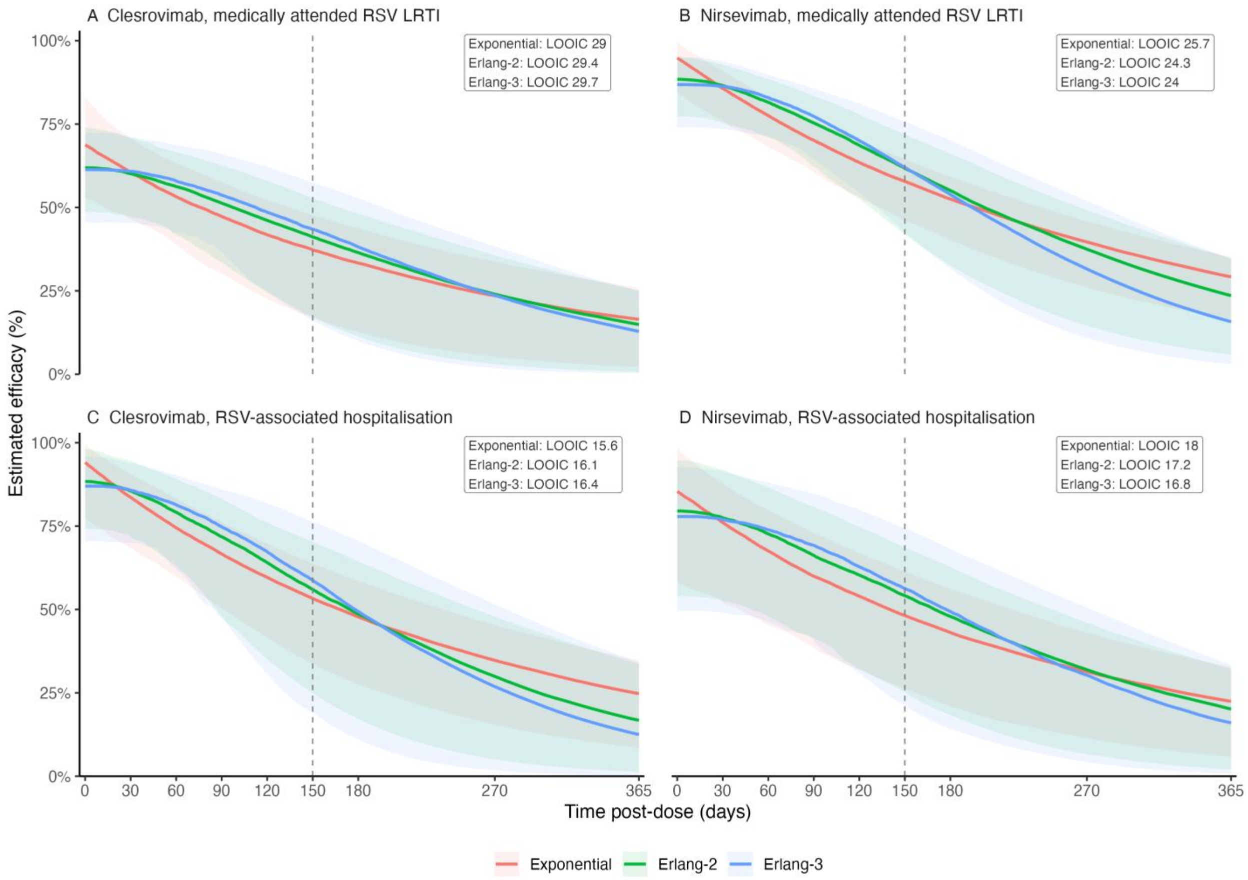


**Supplementary Figure S2.** Sensitivity of estimated waning efficacy to alternative parametric specifications. Model-estimated efficacy over 365 days post-dose using exponential, Erlang-2, and Erlang-3 waning functions for (A) clesrovimab against medically attended RSV LRTI, (B) nirsevimab against medically attended RSV LRTI, (C) clesrovimab against RSV-associated hospitalisation, and (D) nirsevimab against RSV-associated hospitalisation. Solid lines represent posterior median efficacy estimates and shaded areas represent time-specific 95% credible intervals. The Erlang-3 specification was used for the primary analysis.

### *Supplementary Figure S3*

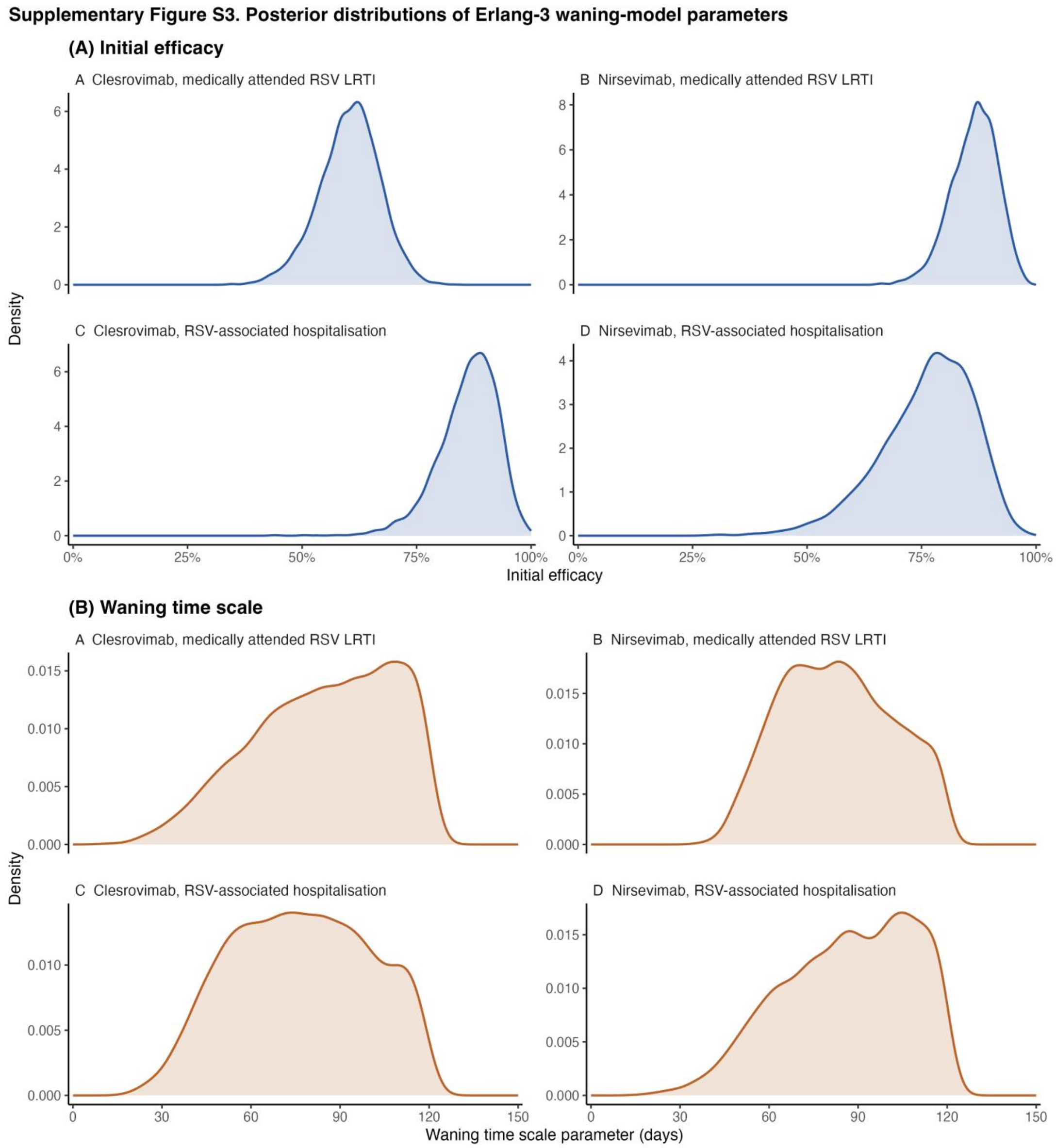


**Supplementary Figure S3.** Posterior distributions of the two key parameters of the primary Erlang-3 waning specification: (A) initial efficacy, and (B) the waning timescale. Within each panel, the four subpanels correspond to (i) clesrovimab against medically attended RSV lower respiratory tract infection (LRTI), (ii) nirsevimab against medically attended RSV LRTI, (iii) clesrovimab against RSV-associated hospitalisation, and (iv) nirsevimab against RSV-associated hospitalisation.

***Supplementary Figure S4***

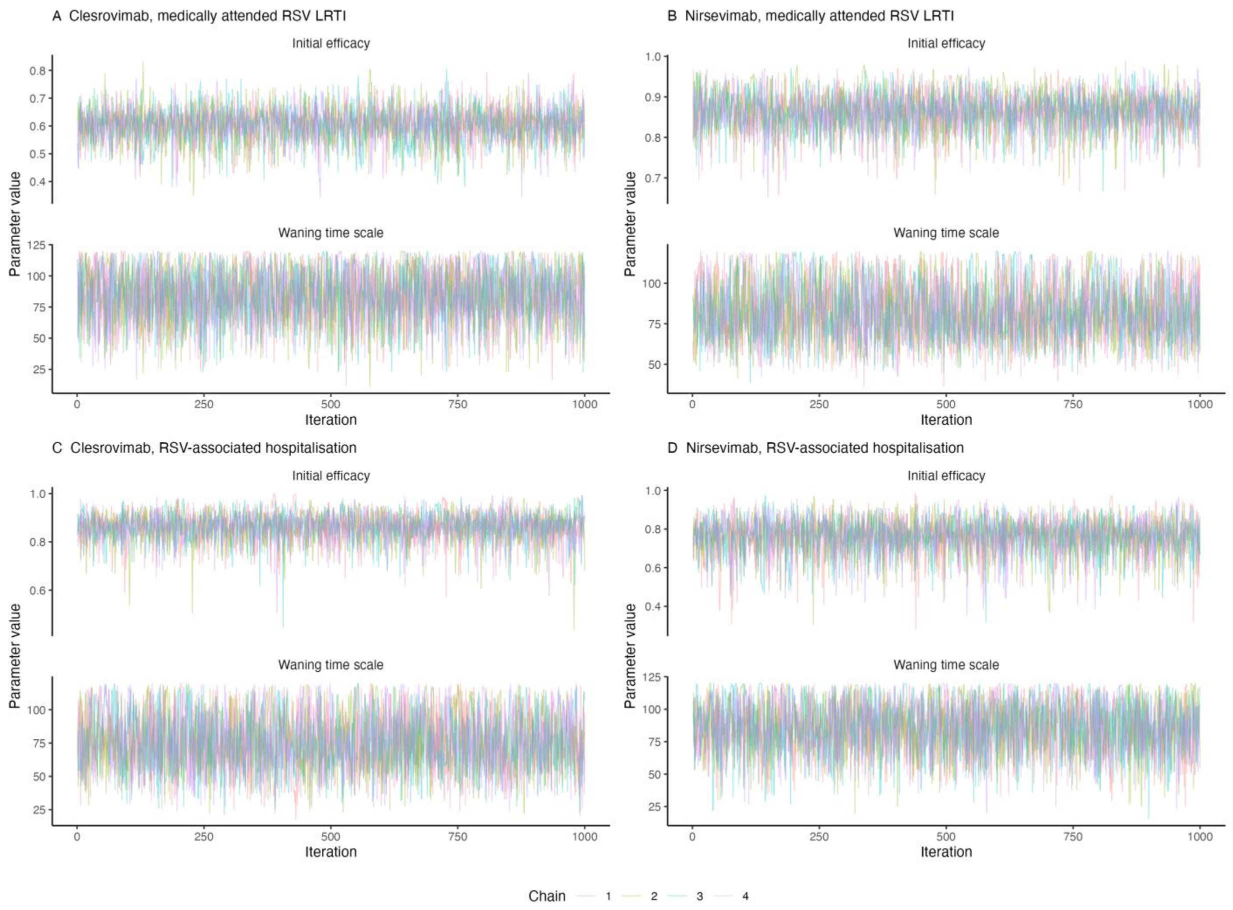


**Supplementary Figure S4.** Markov chain Monte Carlo trace plots of initial efficacy and the waning timescale for the primary Erlang-3 waning specification, shown for four chains across (A) clesrovimab against medically attended RSV LRTI, (B) nirsevimab against medically attended RSV LRTI, (C) clesrovimab against RSV-associated hospitalisation, and (D) nirsevimab against RSV-associated hospitalisation.

***Supplementary Figure S5***

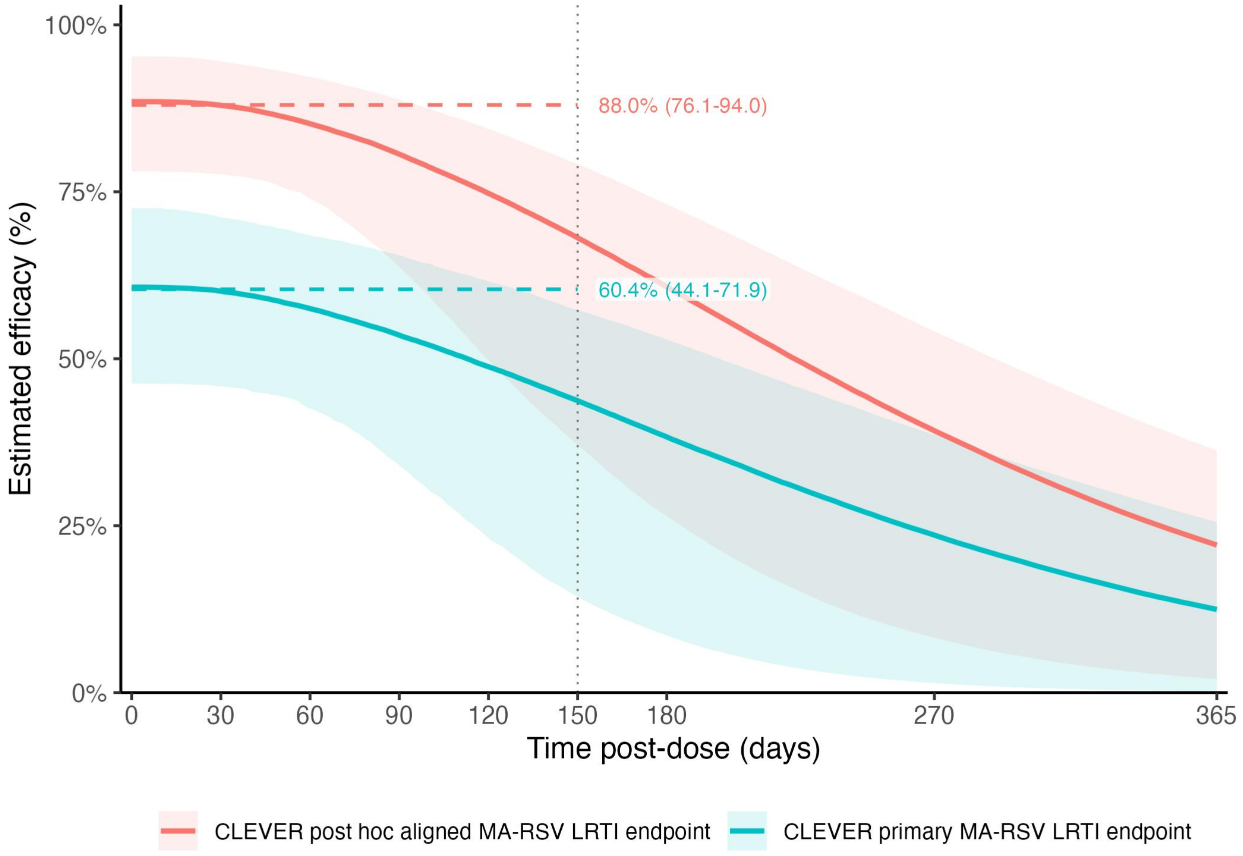


**Supplementary Figure S5.** Exploratory endpoint-alignment sensitivity analysis for clesrovimab against medically attended RSV LRTI. Model-estimated time-specific efficacy under the CLEVER primary endpoint (at least one lower-respiratory-infection indicator **or** at least one disease-severity indicator) and under the CLEVER post hoc endpoint (at least one lower-respiratory-infection indicator **and** at least one disease-severity indicator, more closely aligned with the nirsevimab case definition). Solid lines show posterior medians; shaded areas show 95% credible intervals. Horizontal dashed lines show published 0–150-day cumulative efficacy with 95% confidence intervals. The vertical dotted line marks day 150.

### *Supplementary Table S1*

**Supplementary Table S1.** Leave-one-out information criterion (LOOIC) values for exponential, Erlang-2, and Erlang-3 waning specifications across the four product–endpoint combinations. ΔLOOIC indicates the difference relative to the best-fitting specification within each combination.

| **Scenario** | **Specification** | **LOOIC** | **LOOIC SE** | **ΔLOOIC** |
|---|---|---|---|---|
| A. Clesrovimab, MA RSV LRTI | Exponential | 29.00 | 3.13 | 0.00 |
| | Erlang-2 | 29.44 | 2.97 | 0.44 |
| | Erlang-3 | 29.68 | 2.85 | 0.69 |
| B. Nirsevimab, MA RSV LRTI | Exponential | 25.73 | 6.35 | 1.68 |
| | Erlang-2 | 24.32 | 4.37 | 0.28 |
| | Erlang-3 | 24.05 | 3.86 | 0 |
| C. Clesrovimab, RSV hosp. | Exponential | 15.59 | 1.05 | 0 |
| | Erlang-2 | 16.10 | 1.73 | 0.50 |
| | Erlang-3 | 16.45 | 2.01 | 0.85 |
| D. Nirsevimab, RSV hosp. | Exponential | 17.99 | 2.98 | 1.23 |
| | Erlang-2 | 17.16 | 2.49 | 0.40 |
| | Erlang-3 | 16.76 | 2.41 | 0 |

#### *Supplementary Table S2*

**Supplementary Table S2.** Sensitivity of cumulative efficacy estimates to the upper bound on the waning timescale prior. For each product–endpoint combination, model-derived posterior means and 95% credible intervals are shown for the primary prior (upper bound 120 days) and a relaxed prior (upper bound 365 days) at 0–150 days post-dose.

| **Product** | **Endpoint** | **Primary prior (120 days)** | **Relaxed prior (365 days)** |
|---|---|---|---|
| | | **0–150-day cumulative efficacy** | **0–150-day cumulative efficacy** |
| Clesrovimab | Medically attended RSV LRTI | 57.0 (43.6–68.4) | 58.4 (45.5–69.2) |
| | RSV-associated hospitalisation | 80.5 (65.8–90.6) | 81.7 (67.5–91.7) |
| Nirsevimab | Medically attended RSV LRTI | 80.0 (69.7–88.1) | 80.8 (70.9–88.5) |
| | RSV-associated hospitalisation | 71.2 (48.6–86.9) | 74.3 (53.1–88.4) |
